# Aortic stenosis post-COVID-19: A mathematical model on waiting lists and mortality

**DOI:** 10.1101/2021.11.11.21266212

**Authors:** Christian P Stickels, Ramesh Nadarajah, Chris P Gale, Houyuan Jiang, Kieran J Sharkey, Ben Gibbison, Nick Holliman, Sara Lombardo, Lars Schewe, Matteo Sommacal, Louise Sun, Jonathan Weir-McCall, Katherine Cheema, James H F Rudd, Mamas A. Mamas, Feryal Erhun

**Affiliations:** Department of Mathematical Sciences, University of Liverpool, UK; Leeds Institute for Cardiovascular and Metabolic Medicine, University of Leeds, UK; Leeds Institute of Data Analytics, University of Leeds, UK; Department of Cardiology, Leeds Teaching Hospitals NHS Trust, Leeds, UK; Judge Business School, University of Cambridge, UK; Cardiac Anaesthesia and Intensive Care, Bristol Medical School, University of Bristol, UK; School of Computing, Newcastle University, UK; Mathematical Sciences, Loughborough University, UK; School of Mathematics, University of Edinburgh, UK; Department of Mathematics, Physics and Electrical Engineering, Northumbria University, UK; Division of Cardiac Anaesthesiology, University of Ottawa Heart Institute, Canada; School of Epidemiology and Public Health, University of Ottawa, Canada; Institute for Clinical Evaluative Sciences, Canada; Department of Radiology, University of Cambridge, UK; British Heart Foundation, UK; Division of Cardiovascular Medicine, University of Cambridge, UK; Keele Cardiovascular Research Group, Keele University, UK

## Abstract

**Objectives:** To provide estimates for how different treatment pathways for the management of severe aortic stenosis (AS) may affect NHS England waiting list duration and associated mortality.

**Design:** We constructed a mathematical model of the excess waiting list and found the closed-form analytic solution to that model. From published data, we calculated estimates for how the following strategies may affect the time to clear the backlog of patients waiting for treatment and the associated waiting list mortality.

**Interventions:** 1) increasing the capacity for the treatment of severe AS, 2) converting proportions of cases from surgery to transcatheter aortic valve implantation, and 3) a combination of these two.

**Results:** In a capacitated system, clearing the backlog by returning to pre-COVID-19 capacity is not possible. A conversion rate of 50% would clear the backlog within 666 (95% CI, 533–848) days with 1419 (95% CI, 597–2189) deaths whilst waiting during this time. A 20% capacity increase would require 535 (95% CI, 434–666) days, with an associated mortality of 1172 (95% CI, 466–1859). A combination of converting 40% cases and increasing capacity by 20% would clear the backlog within a year (343 (95% CI, 281–410) days) with 784 (95% CI, 292–1324) deaths whilst awaiting treatment.

**Conclusion:** A strategy change to the management of severe AS is required to reduce the NHS backlog and waiting list deaths during the post-COVID-19 ‘recovery’ period. However, plausible adaptations will still incur a substantial wait and many hundreds dying without treatment.

## Introduction

The COVID-19 pandemic has led to the reorganisation of healthcare services to limit the transmission of the virus and deal with the sequelae of infection. This reorganisation had a detrimental effect on cardiovascular services, with reductions in hospitalisations for acute cardiovascular events and the deferral of all but the most urgent interventional procedures and operations.[1, 2]

Aortic stenosis (AS) is the most common form of valvular heart disease. Once stenosis is severe, symptoms follow and the prognosis is poor, with 50% mortality within two years of symptom onset.[3] Thus, timely treatment is of paramount importance. Surgical aortic valve replacement (SAVR) has historically been the default treatment strategy. However, transcatheter aortic valve implantation (TAVI) has recently emerged as an effective and increasingly utilised option across operative risk strata.[4-8]

There was a large decline in TAVI and SAVR procedural activity to treat severe AS during the COVID-19 pandemic.[9] Between the period March to November 2020, it is estimated that the decrease in activity accounted for 4989 (95% CI. 4020–5959) patients in England with severe AS left untreated by TAVI or SAVR.[9] As we move into an era of ‘living with’ COVID-19, plans must urgently be put in place to best manage the additional waiting list burden for treatment of severe AS.[10]

In this study, we used mathematical methods to examine the extent to which additional capacity to provide treatment of severe AS should be created to clear the backlog and minimise deaths of people on the waiting list.

## Methods

### Study population and assumptions

Data from the UK TAVR registry and NICOR (National Institute for Cardiovascular Outcomes Research) National Adult Cardiac Surgery Audit (NACSA) between 2017 and 2020 have previously been extracted to estimate an excess waiting list size (*W*_O_) of 4989 (95% CI, 4020–5959) patients with severe AS left untreated as of November 2020.[9] In the absence of contemporaneous data on waiting lists and SAVR and TAVI activity, we have taken this number as the excess backlog on which to model solutions. The incidence of AS has not increased over recent years.[11] Therefore, we assumed that the system was in a steady state before the COVID-19 pandemic and without loss of generality defined the steady-state waiting list to be zero. Additionally, we assumed that the normal rate of flow (*f*) of new patients into the waiting list for treatment of severe AS would be maintained upon the commencement of additional operations. Thus, the extra capacity that we model is to clear the excess post-COVID-19 backlog.

We took one-year mortality (*μ*) after the onset of symptoms in severe AS to be 36% (95% CI, 12% – 60%).[12] More recent studies have estimated the one-year mortality to be 51%[5] and 55%, but these included cohorts that were considered inappropriate for SAVR, thus, we considered these estimates unrepresentative of an unselected population with severe AS.[13] The routine capacity for treatment of severe AS was taken from the pre-pandemic period. In 2018/19, the NHS in England performed 7830 SAVR 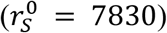 and 5197 TAVI 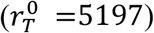 procedures, for a total throughput of about 13,000 per year.[14]

### Modelling

Patients on the waiting list for treatment of severe AS were represented as a dynamical system (figure 1).

**Figure 1:**
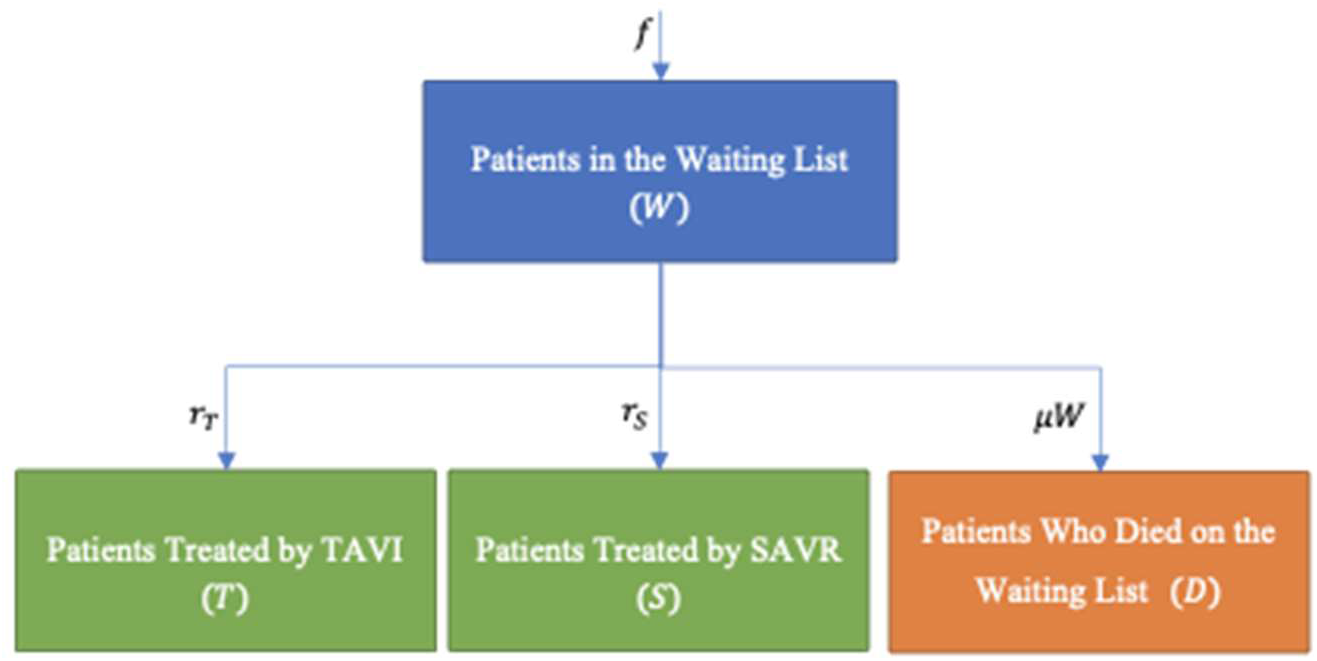
Dynamical system model of the waiting list length

To this model, we introduce capacity in surplus to the 2018/19 performance and call this capacity *T*_e_ (further details are provided in supplementary material). We assume that the typical caseload for which the NHS in England can deal with continues; therefore, the backlog is only reduced by treating patients with this extra capacity or by patient mortality before receiving treatment. We also consider patients in the backlog and patients new to the waiting list indistinguishable. Thus, the waiting list size represents the excess number of people seeking treatment who are unable to be treated immediately at any one time. We also assume that other paths out of the waiting list (i.e. patients seeking private treatment) would be so small in comparison to the uncertainty estimates as to be negligible on the results of our analysis.

These assumptions then come together to give an estimated time (see supplementary material for derivation) to clear the waiting list (*t*_C_)

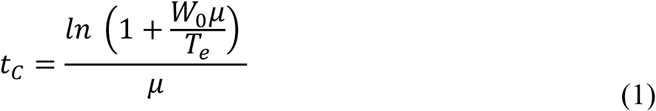

and associated mortality (*m*(*t*_C_))

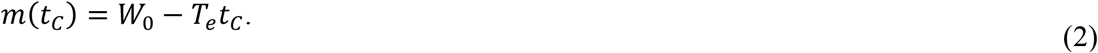

Using equations (1) and (2), we can predict the length of time and associated mortality for different percentage increases in capacity. We assume any capacity increase to be constant throughout the entire modelled period. For example, if we increased daily capacity by 5% this would result in, 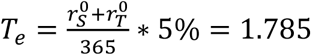 extra procedures per day, across the whole of the NHS in England. We generated 10,000 random values for the one-year mortality rate and initial waiting list length. We assumed that the uncertainty in both variables was normally distributed.

### Interventions and outcomes

We investigated three types of capacity increase: 1) a general increase in the capacity to provide SAVR and TAVI, which could be facilitated by an increased number of procedures per list, additional lists, and prioritisation of care pathways and staffing to treat severe AS; 2) extra capacity created by treating some patients with TAVI who would routinely have SAVR; 3) a combination of a general increase in capacity and the conversion of a proportion of cases from SAVR to TAVI. During the COVID-19 pandemic, TAVI was performed in patients usually referred for surgery, with no difference in short term outcomes compared to historical reference groups.[15, 16]

We assumed that the duration of a SAVR would routinely be between 2-4 hours and a TAVI between 1-2 hours.[17, 18] As such, we assumed within the time for two SAVR operations, three TAVI could be performed instead.[19] Several clinical factors may favour SAVR over TAVI (including concomitant severe coronary artery disease, low STS score, bicuspid aortic valve etc.); therefore, we assumed that, in the short term, no more than 50% of patients could be converted from SAVR to TAVI.[20] We also assumed that no more than 50% extra capacity could be created by other means (e.g. extra lists, more procedures per list). We simulated two principal outcomes based on the creation of additional capacity (*T*_e_): Time to clear the backlog (reduce to zero), Mortality of patients within the excess backlog whilst on the waiting list to be treated.

We completed additional sensitivity analyses for how the conversion of SAVR to TAVI would affect the principal outcomes, including if three SAVR operations could be routinely completed in a day and four to five TAVI procedures per day (presuming increasing uptake of a minimalist TAVI approach without general anaesthesia enabling more rapid procedure time).[21]

## Results

In the pre-COVID-19 period, the routine capacity for treatment of severe AS was set to cover the normal incident rate. That is, clearing the backlog by returning to pre-COVID-19 capacity is not possible. As a result, mortality on the excess waiting list at one year are estimated to be more than 1500, putting a strong emphasis on the need for change.

### Total additional capacity

Figure 2 provides simulations of the time to clear the excess backlog and the mortality of patients on the waiting list based on the amount of total additional capacity, *T*_e_. With a 5% increase in the capacity to provide treatment of severe AS, we estimate it would take 1384 (95% CI, 1025–1994) days to clear the excess backlog, with 2526 (95% CI, 1355–3516) deaths. A 20% increase in total capacity would provide a sharp benefit in clearing the excess backlog within 536 (95% CI, 434–666) days, with an estimate of 1173 (95% CI, 466–1859) deaths. As total capacity increases further, there is a diminishing return in clearing the backlog and avoiding associated mortality; the greater the capacity increase, the fewer lives are saved for every extra increase in capacity. Even if it was possible to double capacity, it would take 131 (95% CI, 126–137) days to clear the backlog and there would be 313 (95% CI, 118–494) deaths on the waiting list.

**Figure 2:**
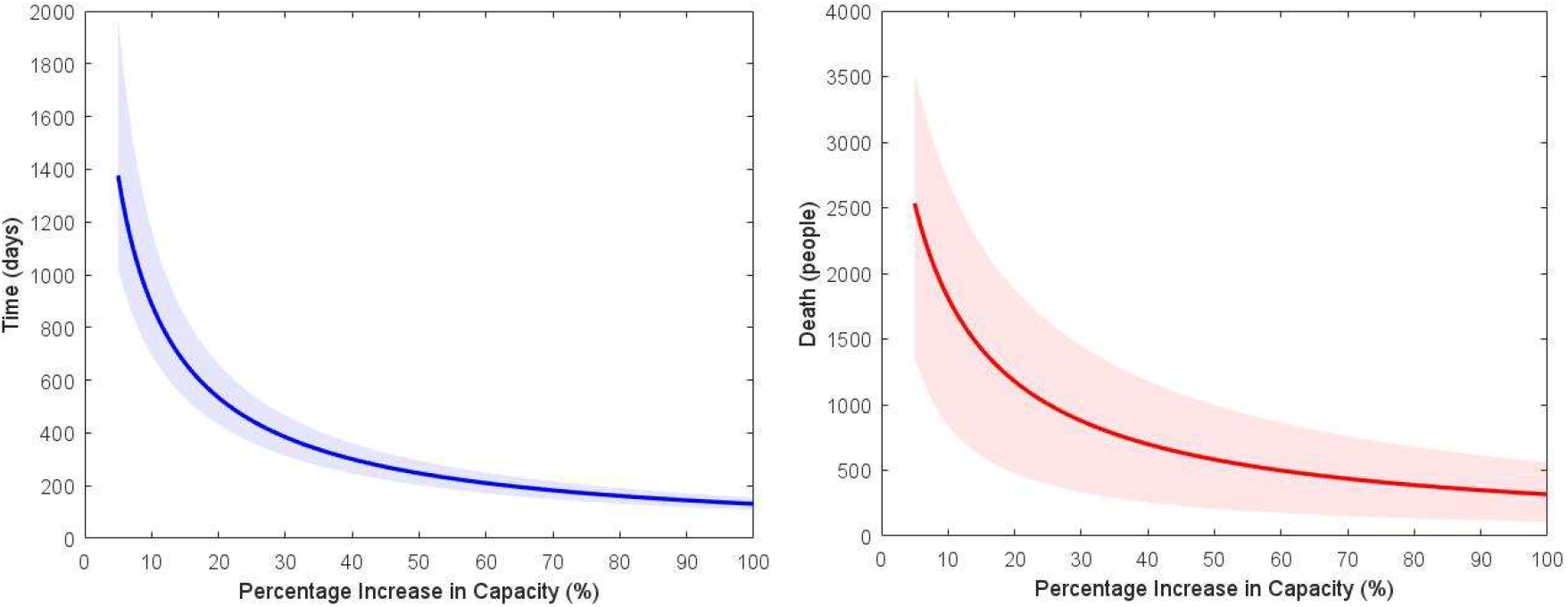
Time to clear backlog (left) and the resulting deaths (right) with associated 95% confidence intervals as a function of daily percentage increase in capacity, with uncertainty from mortality and the initial waiting list. The x-axis is truncated at 5% for visualisation and clarity.

### The effect of converting SAVR to TAVI

The conversion of a proportion of cases from surgery to TAVI provides a modest improvement in estimates of time to clear the backlog and mortality on the waiting list. With the conversion of 30% of SAVR operations to TAVI procedures, without the creation of additional capacity in the system, we estimate it would take 975 (95% CI, 741–1284) days to clear the backlog and result in 1914 (95% CI, 923–2809) deaths on the waiting list. Even with the conversion of 50% of SAVR operations to TAVI procedures, we estimate the backlog would be cleared within 666 (95% CI, 533–848) days with 1419 (95% CI, 597–2189) deaths.

### Combining conversion of SAVR to TAVI and additional capacity

Figures 3a and 3b demonstrate the range of possibilities in creating extra capacity. Each line demonstrates a range of intervention strategies that provide the same result. For example, to reduce mean predicted deaths to 1000 people (red line figure 3b), centres could increase capacity to provide an extra 25% procedures per week at the same mix as pre-pandemic, or they could convert 50% of SAVR operations to TAVI and increase capacity by 8.7% at that mix. Figures 3c and 3d represent how the combinations of interventions to increase capacity within the system alongside the conversion of SAVR to TAVI would impact the time to clear the backlog and on the associated mortality of waiting. Mortality on the waiting list is less responsive to our modelled interventions than the time to clear the backlog (the darker coloured regions of figure 3d make up a greater proportion of the estimates than those of figure 3c). Increasing capacity within the system alongside converting a proportion of SAVR cases to TAVI provides the greatest benefit in clearing the backlog and avoiding associated mortality. A combination that would result in the clearance of the backlog within a year might be of interest for decision makers. With the conversion of 40% of SAVR operations to TAVI and creation of an additional 20% capacity, we estimate the backlog would be cleared in just under a year – 343 days (95% CI, 281–410) with 784 (95% CI, 292–1324) deaths before treatment. Sensitivity analyses where the number of TAVI procedures that could be completed within the same time as SAVR was altered (TAVI to SAVR: 4 to 3, 4 to 2, 5 to 3) support these findings (supplementary material figures S1 – S3). Furthermore, sensitivity analyses show that with the best-in-class practices (TAVI to SAVR: 4 to 2), even a more modest combination (a conversion of 35% and creation of an additional 10% capacity) would be enough to clear the backlog within a year.

**Figure 3:**
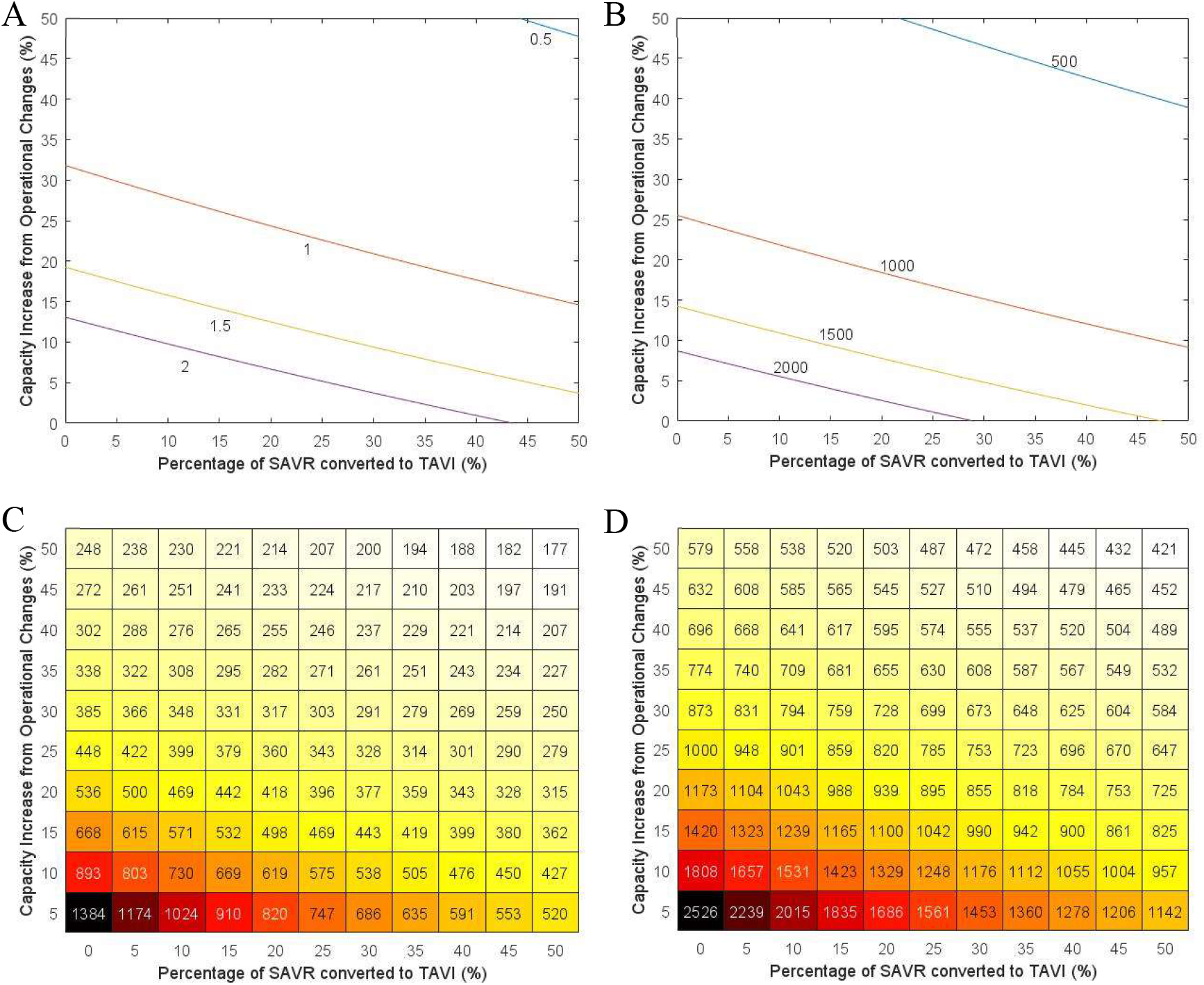
Mean time to clear backlog (left) and the resulting deaths (right) as a function of daily percentage increase in capacity (y-axis) and percentage of SAVR converted to TAVI (x-axis) (Presented in two different forms). A) Isoclines of constant mean clearance-time going from half a year (blue) to 2 years (purple) in half-year increments. B) Isoclines of constant mean mortality after clearing the backlog from 500 people (blue) to 2000 (purple) in 500-person increments. C) Heatmap of different combinations of conversion and daily capacity increases and how long the backlog would take to clear on average, in days. D) Heatmap of different combinations of conversion and daily capacity increases and how many people would die, on average.

## Discussion

In this study, using dynamical system modelling, we provide estimates for how changes to treatment pathways for severe aortic stenosis may affect the time taken to clear the backlog and minimise mortality on the waiting list in the NHS of England. Without providing at least 20% total additional capacity for the interventional treatment of AS, we estimated there would be more than 1000 deaths on the waiting list over a period of nearly 1.5 years. A conversion of cases from SAVR to TAVI would expedite the clearance of the backlog, but even converting half the cases to TAVI would still result in over 1400 deaths over a period of almost 2 years. A combination of converting 40% of cases usually planned for SAVR to TAVI and creating 20% additional capacity for procedures (through measures such as extra lists) would clear the excess backlog within one year, with 784 deaths.

Our study has several strengths. First, in an urgent situation of many unknowns, our use of novel mathematical models provides plausible estimates on which to base planning and provides an exemplar that may be used in service delivery in other conditions in the post-pandemic landscape. Given the high event rate amongst this population, waiting for more contemporary data to be collected may well not provide enough time to institute system changes to prevent deaths. Second, we also provide specific estimates for how the conversion of cases to TAVI from surgery may affect waiting lists and associated mortality, which can inform local MDT discussions. Third, our model can act as a basis for a clinical and cost-benefit analysis to evaluate different ways to increase capacity and define the optimal strategy at each centre. For each centre, the most effective combination of converting SAVR to TAVI and provision or prioritisation of treatment of severe AS can be generated.

We also recognise the limitations inherent in modelling a complex situation. First, we represent the NHS in England as a single entity. As such, we implicitly assume that population and capacity are distributed evenly throughout the country by treating centre capacity. If the distribution of waiting list patients deviates significantly from the distribution of treatment centres weighted by capacity, the time it would take to clear the waiting list, and thus the mortality rate would be higher. Second, we have not attempted to calculate how many AS patients may have died in the COVID-19 pandemic. Third, our assumed mortality rate may differ at a centre-level due to prioritising clinically more vulnerable patients on the waiting list. Fourth, a centre-level analysis could account for the different practices in each treatment centre and identify strategies that work best for each centre. Fifth, our estimates from converting cases from SAVR to TAVI does not include post-procedural factors such as the requirement for intensive care capacity, hospital stay and further procedures because these rely on multiple centre-specific factors. Finally, it has been shown that rapid growth in the demand for TAVI can overwhelm current capacity,[22] which may lead to prolonged wait times and subsequent adverse outcomes while patients are on the waitlist. Therefore, a demand model that captures the changes of demand for TAVI and SAVR would be a helpful future direction of analysis.

A previous study used a mathematical model to quantify the cumulative cardiac surgical backlog (including coronary artery bypass grafting surgery, valve replacement and transcatheter aortic and mitral valve replacements) in two centres based on the projected pandemic duration in the United States of America (USA).[23] The authors used simple mathematical models to predict the time required to clear the backlog depending on increased operating capacity. However, the authors did not consider mortality, which we have as it is of critical importance to patients and when planning services.

The results of our study highlight concerns pertaining to the deferral of non-emergency treatment for severe AS during the ‘recovery period’ of COVID-19. Severe AS is a progressive condition with valve replacement the only available treatment improving prognosis.[24] On a local, regional, and national scale, healthcare systems will need to examine capacity, set priorities, and plan for adequate capacity to manage the backlog of patients with severe AS. The response will be complicated by prior exhaustion of human resources from the pandemic and competition with other specialities, which will also have backlogs.[25]

Nonetheless, planning should prioritise patients at the highest risk from a deferral of treatment. Mortality on the waiting list for AS has been reported to be as high as 14%.[26] Furthermore, patients awaiting structural procedures deferred due to the pandemic have been found to have significantly higher mortality rates compared to those with stable coronary artery disease.[27] Prioritising capacity for treatment of patients with severe AS may mean reduced capacity for other procedures. This interaction will require collaborative decision-making on a local level accepting that these are difficult, imperfect times. We also show that the conversion of a proportion of cases that would usually be managed by SAVR to TAVI can help expedite treatment and reduce mortality on the waiting list. During the pandemic, TAVI procedures were performed in patients usually referred for surgery with no apparent difference in short term outcomes;[15, 16] and data continues to emerge for longer-term efficacy and safety of TAVI across operative risk strata.[28,29] Recent European guidelines suggest that TAVI would be a preferable option for patients over 75 years of age compared to SAVR.[20] To help planning, we provide an app (https://github.com/Christian-P-Stickels/AS_Waitinglist_data) to explore the impact of alterations in capacity and treatment pathways on waiting lists and mortality related to severe AS at a local, regional and national level (supplementary material).

### Conclusions

In this study, we identify that without a combination of increased capacity for treatment of patients with severe aortic stenosis, and consideration of expanding the use of TAVI, there will be unpalatable rates of mortality in this high-risk group during the post-COVID-19 ‘recovery’ period. These results should inform the planning of cardiac services.

## Data Availability

This manuscript does not present new experimental data.

## Acknowledgements

We want to thank all the participants of the V-KEMS Study Group on “Modelling Solutions to the Impact of COVID-19 on Cardiovascular Waiting Lists” that took place on February 2-4, 2021, for thought-provoking discussions. Our special thanks to Clare Merritt (Newton Gateway to Mathematics), whose help extended beyond the workshop and was crucial in completing this work, and to Alan Champneys who brought the group together in the first place.

## Supplement

### Supplement 1: Mathematical Derivation of the Differential Equation and its Solution

From figure 1, we can write the following equation:

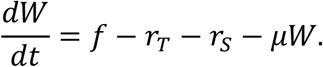

We can then re-write and integrate this equation

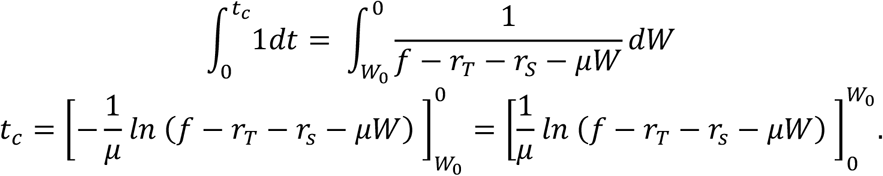

We can now define *T*_e_, the extra capacity, as *T*_e_ = *r*_T_ + *r*_s_ − *f*. This is because we claim that under normal conditions, 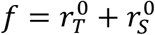, such that the waiting list never grows above zero, and that the additional patients are already on the waiting list. The equation for *T*_e_ follows the observation that the current rates of TAVI and SAVR treatment are the normal rates plus the additional capacity.

This substitution allows us to write

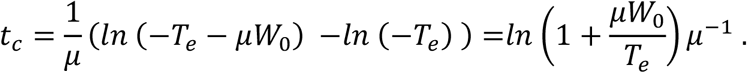

This is the solution we use for calculating the time when the waiting list becomes zero.

We now rely on the assumption that *T*_e_ is constant to write

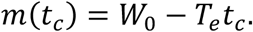

That is, by the time the waiting list is zero, everyone who has not been treated is unfortunately dead.

The assumption of a front-loaded waiting list (i.e., that all additional patients are identified and waiting) is not a strict requirement for this model to be valid. If it is the case that the additional patients are still being identified when the extra capacity is created, then as long as they are identified at a faster rate than they are treated, the predictions in this model hold. It is only in cases where the identification rate is less than the treatment rate that this assumption becomes invalid. In such cases, *T*_e_ can be said to be equal to the identification rate instead.

This is true because mortality is not tied to being on the waiting list but from the onset of symptoms. In this way, the waiting list in our model can be thought of as the list of all people who need treatment, even if the NHS is unaware of them.

This model can be extended to predict mortality and time to clear a waiting list for non-constant *T*_e_, but we do not expand on that here.

### Supplement 2: Data

We calculate the increase in capacity due to conversions and operational changes as follows. Assume that we increase operations by 20% due to operational changes and convert 10% of all SAVR to TAVI. Also assume that for every three SAVR patients five TAVI patients can be processed. If we convert 10% of SAVR cases to TAVI (783 SAVR patients), we can treat an additional 522 patients from the waiting list. From the 20% increase, we get extra 1039 TAVI and 1566 SAVR operations per year. If we apply 10% conversion to this extra capacity, 156 SAVR operations can be converted into 260 TAVI operations. In total, the operational changes and conversion create an extra capacity of 3232 operations with which to service the waiting list each year: 1822 (1,039+522+261) TAVI and 1410 (1,566-156) SAVR operations.

N.B. We make no assumptions about who the extra TAVI procedures treat, for example, if in the above example, the additional 626 TAVI procedures we gain from conversion (522 from converting the normal capacity and 104 from converting the additional capacity) treated only SAVR patients, the conversion rate would actually be 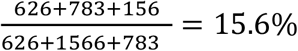;. Normally, we would expect that the application of this extra TAVI would be in the same proportion as the ratio of SAVR to TAVI, which would give a real-world conversion rate of 13.5%.

### Supplement 3: App

The app can be accessed at https://github.com/Christian-P-Stickels/AS_Waitinglist_data

### Supplement 4: Additional Results

**Supplementary figure S1:**
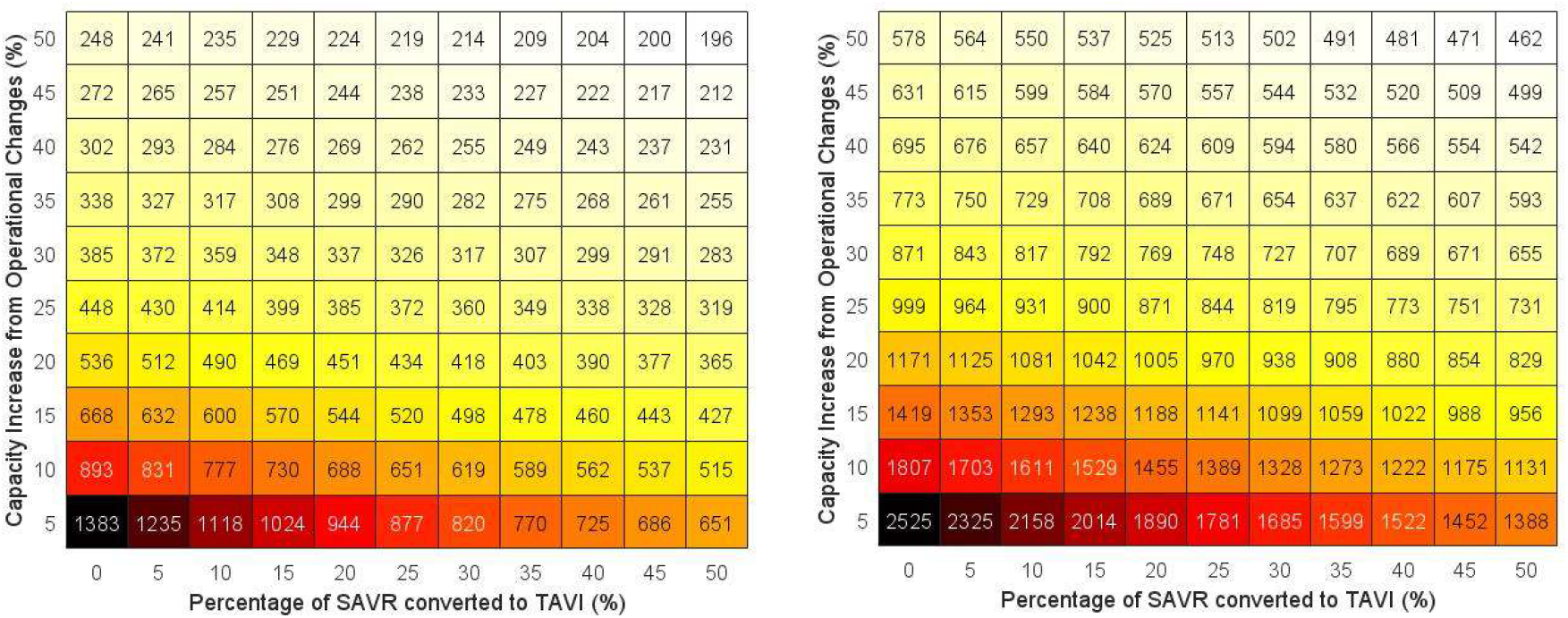
Heat map of a three-to-four SAVR-to-TAVI conversion. Mean time to clear backlog (left) and the resulting deaths (right) as a function of daily percentage increase in capacity (y-axis) and percentage of SAVR converted to TAVI (x-axis), assuming that for every three SAVR operations, four TAVI procedures can be performed instead.

**Supplementary figure S2:**
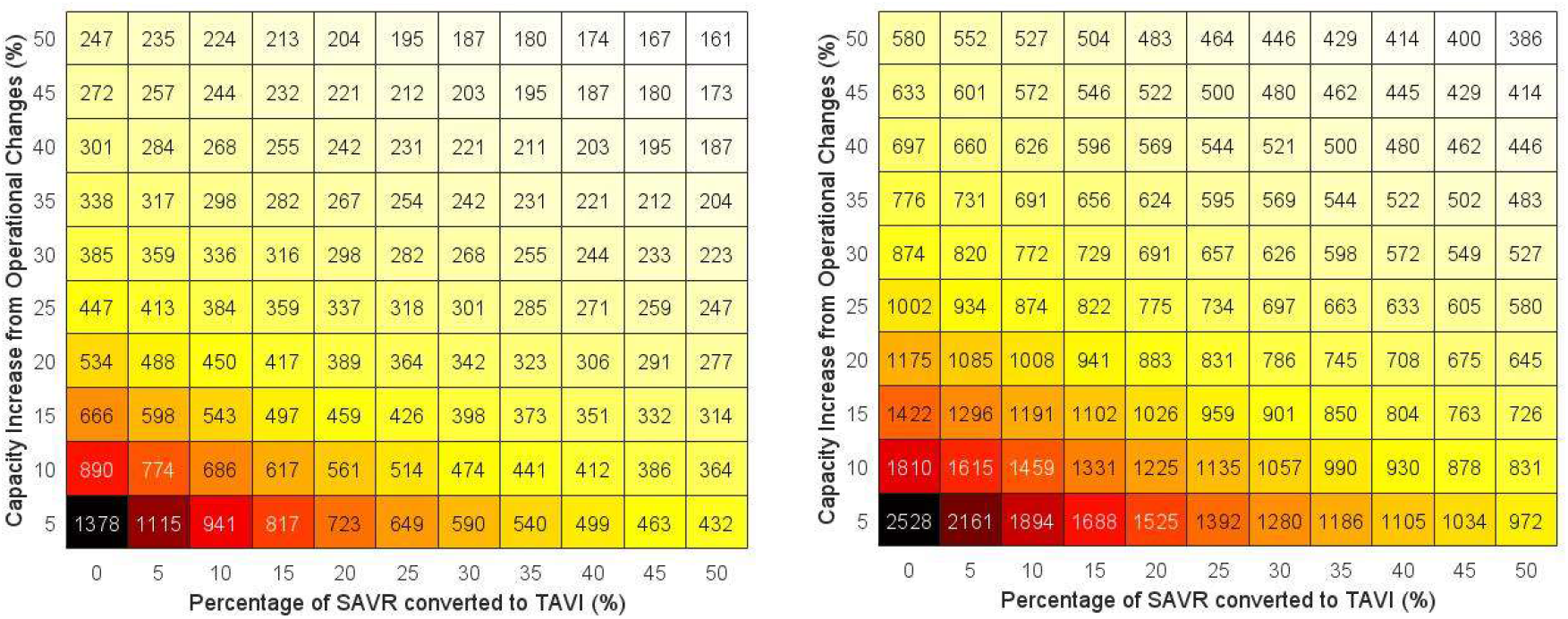
Heat map of a three-to-five SAVR-to-TAVI conversion. Mean time to clear backlog (left) and the resulting deaths (right) as a function of daily percentage increase in capacity (y-axis) and percentage of SAVR converted to TAVI (x-axis), assuming that for every three SAVR operations, five TAVI procedures can be performed instead.

**Supplementary figure S3:**
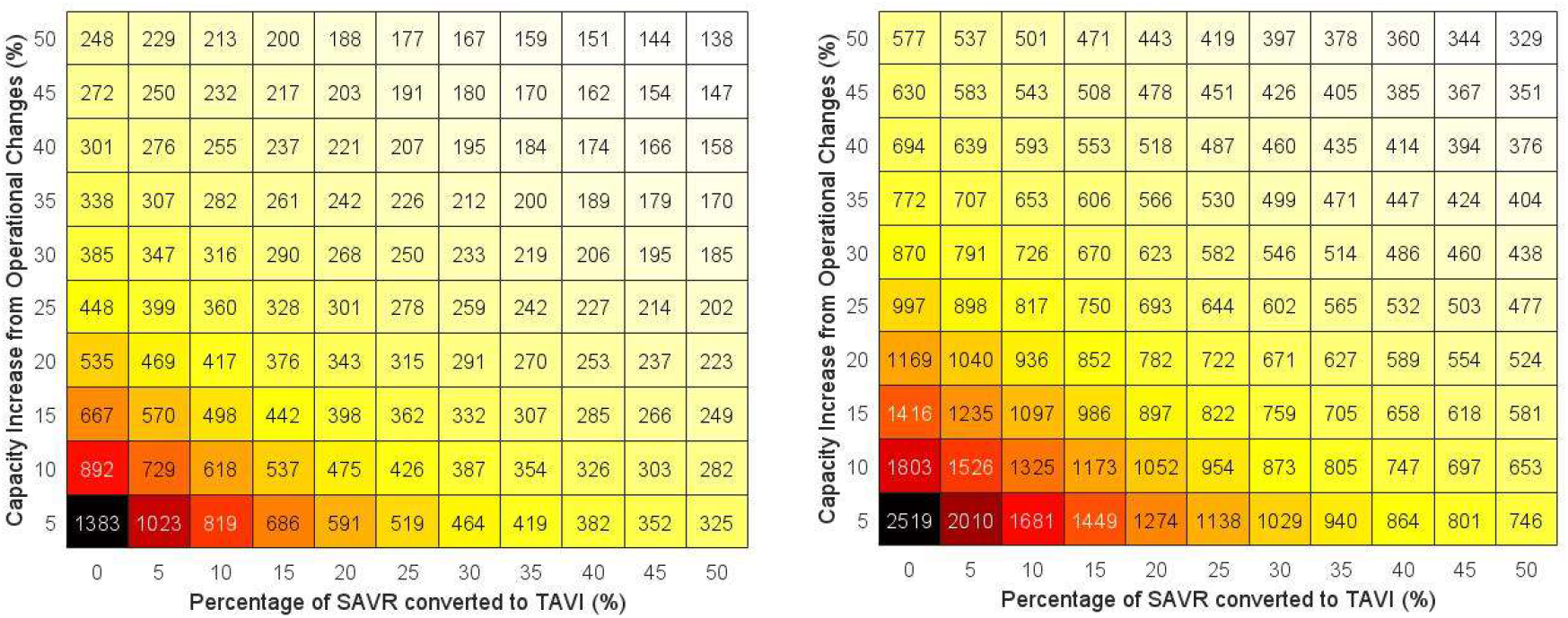
Heat map of a two-to-four SAVR-to-TAVI conversion. Mean time to clear backlog (left) and the resulting deaths (right) as a function of daily percentage increase in capacity (y-axis) and percentage of SAVR converted to TAVI (x-axis), assuming that for every two SAVR operations, four TAVI procedures can be performed instead.

**Supplementary figure S4:**
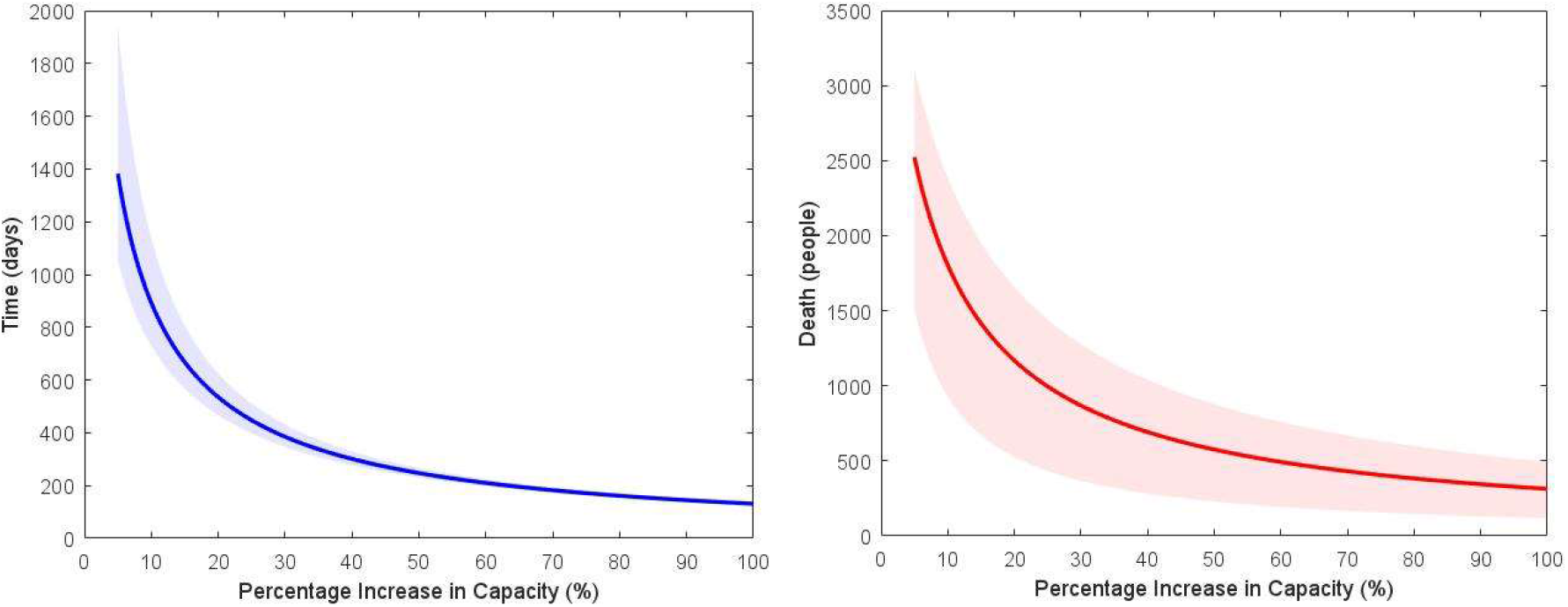
Error from mortality estimates. Time to clear backlog (left) and the resulting deaths (right) with associated 95% confidence intervals as a function of daily percentage increase in capacity, with uncertainty from mortality only. The x-axis is truncated at 5% for visualisation and clarity.

We find that error in the one-year mortality causes higher uncertainty at lower capacity increases, but at higher capacity increases, this uncertainty decreases until it is almost zero with regards to clearance time. This is likely because at higher capacity increases, more of our waiting list clearance comes from treatment, as opposed to death, resulting in less error.

**Supplementary figure S5:**
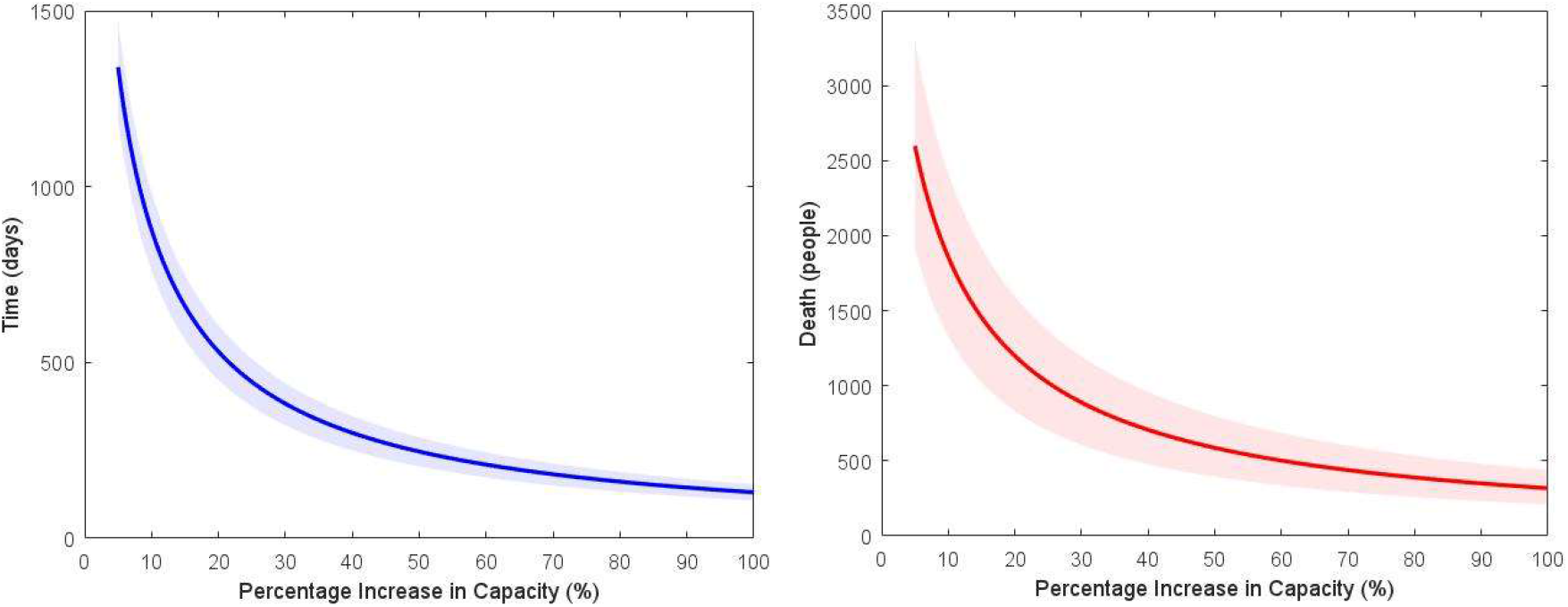
Error from wait list (W0) estimates. Time to clear backlog (left) and the resulting deaths (right) with associated 95% confidence intervals as a function of daily percentage increase in capacity, with uncertainty from initial waiting list estimates only. The x-axis is truncated at 5% for visualisation and clarity.

We find that error in the estimate of the wait list length W0 causes uncertainty that is fairly constant in the time it takes to clear the backlog and in resultant deaths. This is to be expected as we can show that the uncertainty scales with ln *W*_O_. There is a small decrease in uncertainty as we increase capacity, once again because an increase in capacity results in more control of the waiting list reduction.

